# Effect of cell isolation magnetic particles on DNA quantification by UV absorbance spectrophotometry

**DOI:** 10.1101/2023.04.24.23288526

**Authors:** Izza Usman Bajwa, Samuel Sigaud

## Abstract

Magnetic particles are commonly used to isolate specific cell types from blood samples. Residual particles present in the genomic DNA extracted from these cells can interfere with concentration measurements by UV absorbance spectrophotometry. In this study, we determined the degree of inaccuracy of DNA quantification by UV spectrophotometry, in the context of the lineage-specific chimerism analysis workflow. We showed that the presence of residual magnetic particles and RNA leads to an overestimation of the DNA concentration.

## Introduction

The use of magnetic particles to isolate specific cell types from blood samples is a common technique in diagnostics or immunogenetics laboratories. For example, the typical workflow for lineage-specific chimerism analysis involves isolating T-Lymphocytes, Myeloid Cells, or other cell types from blood samples, followed by genomic DNA extraction, prior to performing PCR or qPCR ^1^. DNA concentration and quality is typically checked following extraction, to ensure that the downstream PCR reactions are performed in optimal conditions. Depending on the DNA extraction method, residual magnetic particles used for the cell isolation step can be found in the final DNA sample. While these particles generally do not interfere with the subsequent PCR reactions, they could have an influence on the DNA quantification step.

UV absorbance spectrophotometry is the most widely used method to assess DNA concentration and purity. It is fast and does not require the use of standard curves or special reagents. It uses very small amounts of DNA, especially when performed with a cuvette-less spectrophotometer such as the NanoDrop instruments (ThermoFisher Scientific). However, UV absorbance is not selective for DNA ^2^. Concentration measurements can be affected by the presence of contaminants, such as RNA, proteins, chemicals used during DNA extraction, or the magnetic particles used for cell isolation.

To overcome these issues, fluorescent DNA-binding dyes have been developed ^3^. These compounds increase considerably their fluorescence when bound to double-stranded DNA. They have a high specificity and sensitivity, and are now considered the gold standard for DNA quantification. However, as compared to UV spectrophotometry, fluorescence measurements are more time-consuming, require the use of expensive reagents, and the realisation of a DNA standard curve. For these reasons, UV spectrophotometry is still the method of choice to determine DNA concentration when many samples need to be processed rapidly, such as in molecular diagnostics laboratories.

The goal of this study was to determine the degree of inaccuracy of DNA quantification by UV spectrophotometry, in the context of the lineage-specific chimerism analysis workflow. We examined the influence of residual magnetic particles on DNA concentration and quality measurements, and provide recommendations to increase measurement accuracy.

## Materials and Methods

### Samples

Human peripheral whole blood was collected from healthy donors using Institutional Review Board (IRB)-approved consent forms and protocols. The samples were received from the collection site in Vacutainer tubes with acid citrate dextrose, solution A, as anticoagulant (Becton, Dickinson and Company, NJ, USA). The blood was processed within 24 hours after collection.

### Cell isolation

CD66b^+^ or CD3^+^ leukocytes were directly isolated from whole blood by positive selection with magnetic particles (EasySep HLA Chimerism Whole Blood CD66b or CD3 Positive Selection Kits, StemCell Technologies, Vancouver, BC, Canada), following manufacturer’s instructions.

### DNA extraction

DNA extraction from the isolated cells was performed using the QIAamp DNA Blood Mini Kit (Qiagen, Hilden, Germany), as per manufacturer’s instruction for spin protocol, and the extracted DNA was eluted in 200 μL of the Elution Buffer provided with the kit.

### DNA quantification by UV spectrophotometry

A NanoDrop 1000 Spectrophotometer (ThermoFisher Scientific, Wilmington, DE, USA) was used for measuring DNA concentration and quality as per manufacturer’s instructions, using 2 μL of extracted DNA sample.

### DNA and RNA quantification by fluorometry

DNA concentration was determined with the QuantiFluor ONE dsDNA System (Promega, Madison, WI, USA) as per manufacturer’s instructions.

RNA concentration was determined using the Qubit RNA HS Assay Kit (ThermoFisher Scientific, Wilmington, DE, USA) as per manufacturer’s instructions.

Measurements were performed on a Qubit Fluorometer (Thermo Fisher Scientific, Wilmington, DE, USA).

## Results

### Absorbance spectrum of magnetic particles

We first measured the absorbance spectrum of the magnetic particles, diluted in elution buffer, between 220 nm and 350 nm (Fig 1). The maximum absorbance was observed at 220 nm, then decreased rapidly and reached zero at 340 nm. While lower than at 220 nm, the absorbance at 260 nm, the wavelength used for DNA concentration measurements, was still significant.

**Fig 1.**
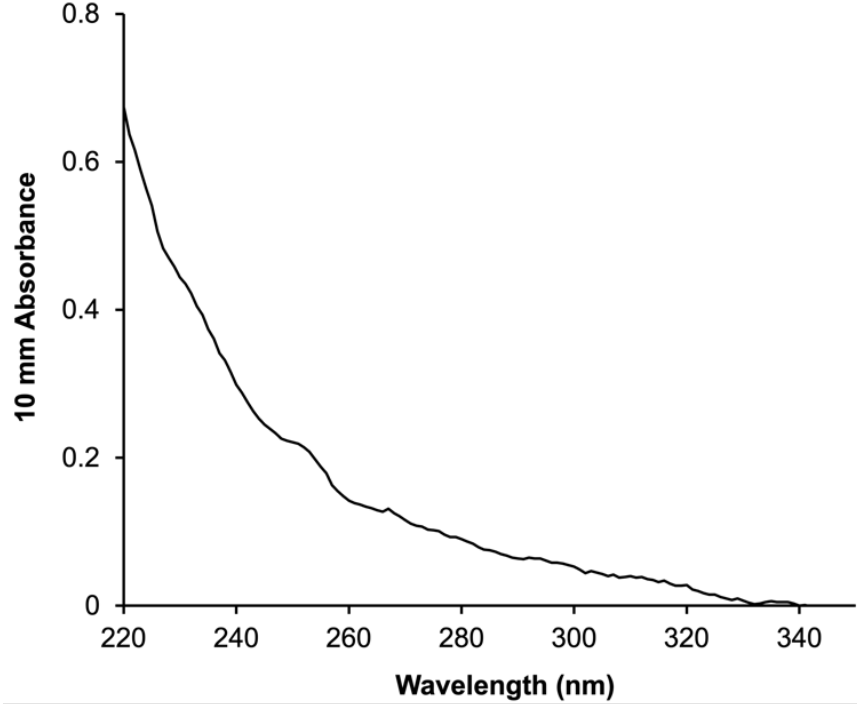
Absorbance spectrum of the magnetic particles.

A standard curve was performed at 220 nm, diluting various quantities of magnetic particles in 200 μL of elution buffer, the quantity used in the standard DNA isolation protocol (Fig 2).

**Fig 2:**
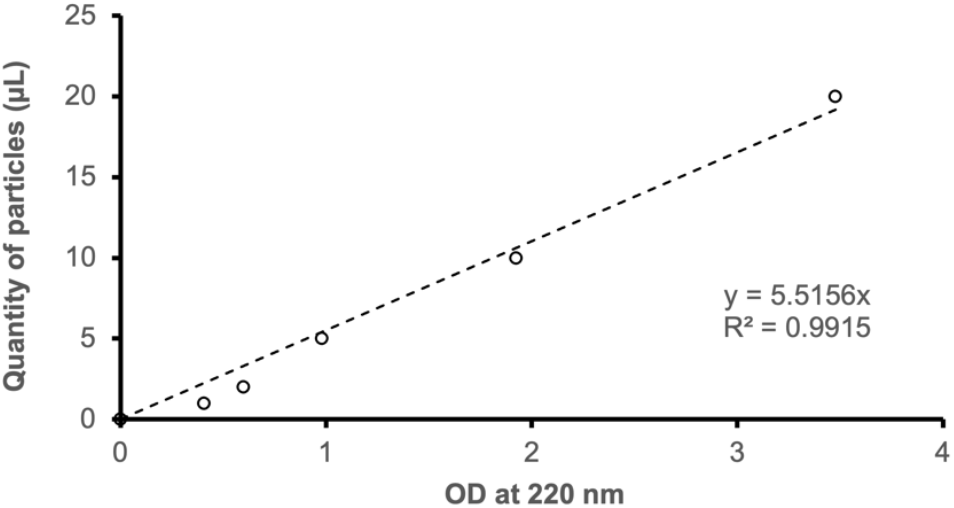
Magnetic particles standard curve at 220 nm. Between 1 and 20 μL of particles were diluted in 200 μL of Elution Buffer, and the absorbance measured at 220 nm with a NanoDrop instrument.

### Quantification of residual magnetic particles

We then aimed to estimate the amount of magnetic particles that were transferred from the initial cell suspensions to the DNA samples. 225 μL of magnetic particles, the amount used for a typical cell isolation from a 3 mL blood sample, were processed through the standard DNA isolation protocol using Qiagen columns. The absorbance at 220 nm of the resulting 200 μL elution was measured and compared to the standard curve established previously. The measured OD (average 0.63) corresponded to 3.47 μL of particles, or about 1.5% of the initial amount of magnetic particles used for cell isolation.

The absorbance at 260 nm of the particle suspensions was on average 0.14, which, if it was present in a DNA sample, would increase the measured DNA concentration by 7 ng/μL.

### Effect of magnetic particles on UV DNA quantification

We then attempted to confirm that the presence of magnetic particles in the DNA samples could influence DNA quantification in standard laboratory settings. CD3^+^ and CD66b^+^ cells were isolated from blood samples from three unrelated donors, and the DNA extracted (samples S1 to S6). DNA concentration was measured by UV absorbance spectrophotometry, before and after removing the magnetic particles by centrifugation. We also measured DNA concentration using a DNA-specific fluorescent dye, which in theory should not be affected by the presence of particles (see Table 1 in data supplements). As expected, magnetic particles contributed significantly to UV absorbance at 260 nm, resulting in an overestimation of the DNA concentration (Fig 3). On average, across the six DNA samples, the calculated DNA concentration decreased by 37% after the magnetic particles were removed from the DNA sample. In contrast, the DNA concentration measured using a fluorescent dye was virtually unchanged after removing the particles.

**Fig 3.**
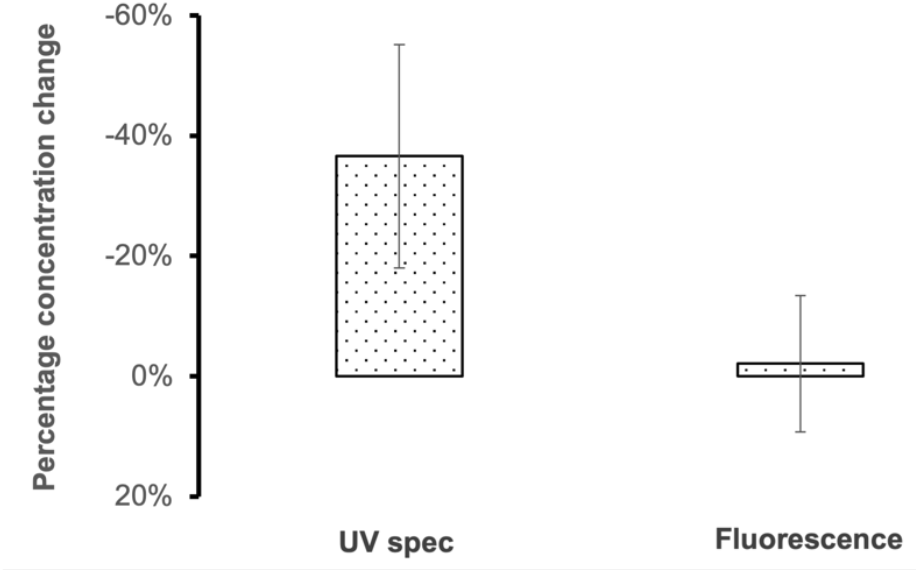
Change in DNA concentration before and after particle removal. DNA concentration was measured after elution from the extraction column, both by UV spectrophotometry and fluorescence. Residual magnetic particles were then removed from the DNA samples by centrifugation, before DNA concentration was measured again. Bars represents standard deviation.

Because of its effect on the absorbance at 260 nm, the presence of magnetic particles in the DNA samples was likely to also influence DNA purity measurements using the A_260_/A_280_ and A_260_/A_230_ ratios. We measured the A_260_/A_280_ ratio (Fig 4A), and the A_260_/A_230_ ratio (Fig 4B) of the DNA samples before and after particle removal by centrifugation. Although the effect was variable depending on the sample, overall removing the particles increased both ratios. The effect was more consistent with the A_260_/A_230_ ratio.

**Fig 4.**
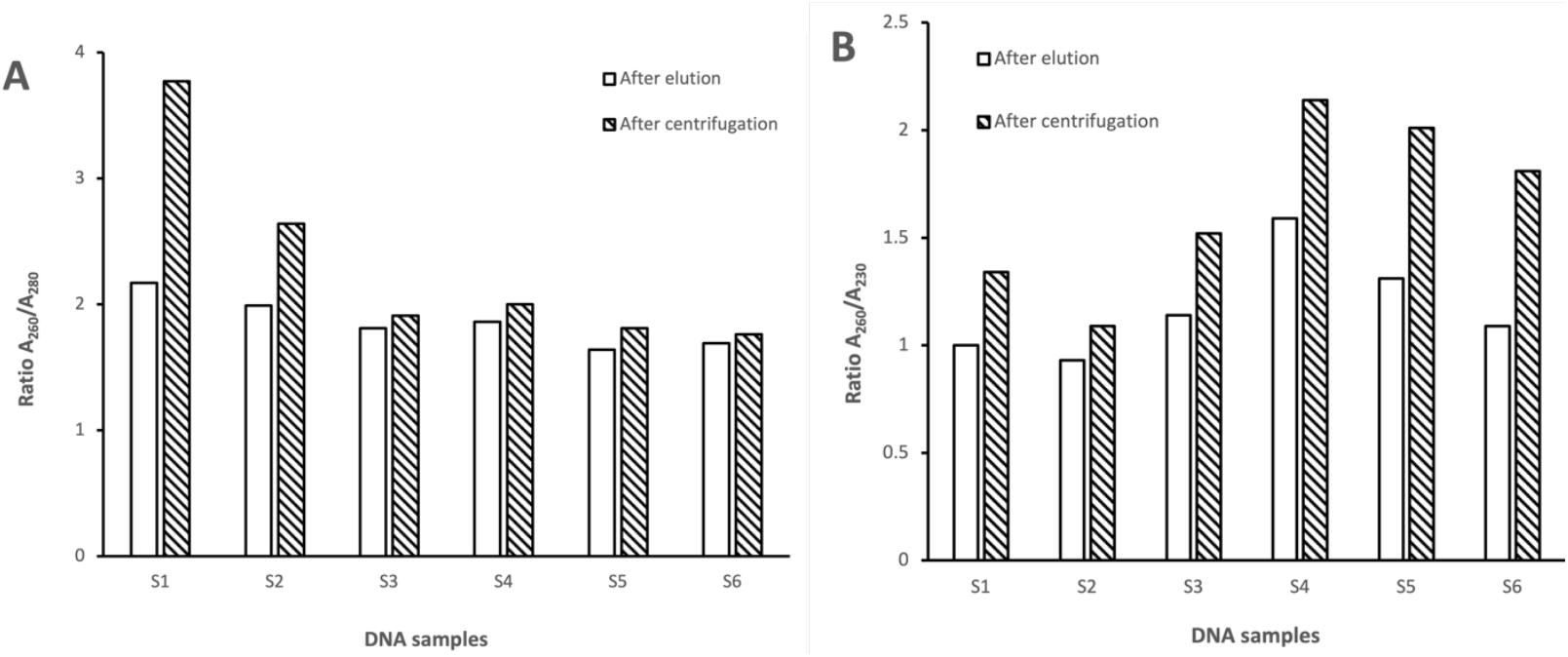
Absorbance ratios before and after particle removal. The absorbance ratios A_260_/A_280_ (A) and A_260_/A_230_ (B) of the DNA samples were measured before and after magnetic particles removal by centrifugation.

### Comparison between UV absorbance spectrophotometry and DNA-specific fluorometric measurements

As we have shown above, removing the particles reduced considerably the DNA concentration when measured by UV absorbance spectrophotometry, while it had no influence on fluorometric measurements. We compared the DNA concentrations measured using the two methods, with and without the presence of magnetic particles (Fig 5). As expected based on the results presented earlier in Fig 3, in the presence of particles, UV measurements overestimated DNA concentration by about 50% as compared to fluorescence measurements. When the particles were removed, the difference in measured DNA concentration was reduced, but was still 20% higher when using UV fluorescence. This suggested that contaminants absorbing at 260 nm, other than the magnetic particles, were present in the DNA samples.

**Fig 5.**
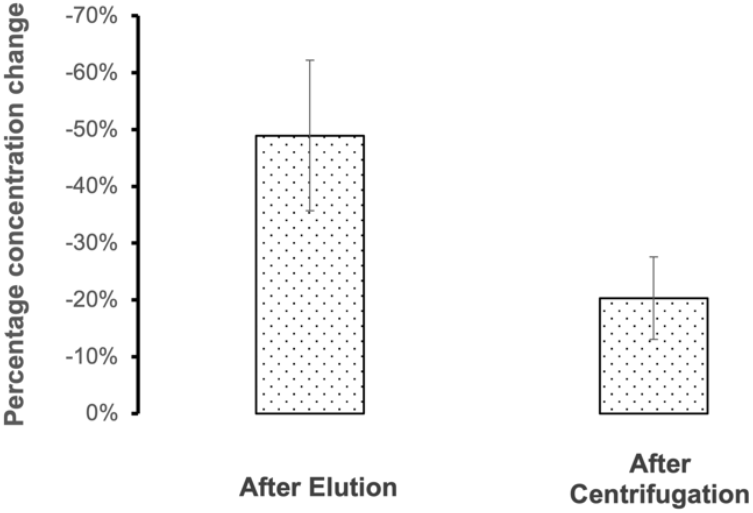
Difference between the DNA concentration measured by UV spectrophotometry and fluorescence. DNA concentration was measured after elution from the extraction column, both by UV spectrophotometry and fluorescence. Residual magnetic particles were then removed from the DNA samples by centrifugation, before DNA concentration was measured again. The difference between the concentration measured by UV spectrophotometry and fluorescence is displayed as a percentage change. Bars represents standard deviation.

Since the DNA extraction method used was not highly specific for DNA^4^, we suspected that most of the difference between UV and fluorescence measurements was due to the presence of RNA in the DNA sample. Indeed, RNA absorbs strongly at 260 nm. In contrast, the fluorescent dye used in our measurements is designed to be highly specific for double-stranded DNA. We used an RNA-specific fluorescent dye to assess the RNA concentration in each of the DNA samples (see Table 1 in data supplements).

We hypothesized that most of the difference between UV absorbance and fluorescence measurement of the DNA concentration was due to the presence of residual magnetic particles and RNA in the genomic DNA samples. To test this hypothesis, for each sample, we added the DNA concentration measured by DNA-specific fluorescence, the contribution to the DNA concentration due to the presence of RNA (measured by RNA-specific fluorescence), and the contribution to the DNA concentration due to the presence of particles (difference between the DNA concentration measured by UV absorbance with and without magnetic particles) (Fig 6). For each sample, the discrepancy in DNA concentration measurements between the two methods (UV absorbance spectrophotometry and DNA-specific fluorescence) could be explained in totality by the presence of residual particles and contaminating RNA.

**Fig 6.**
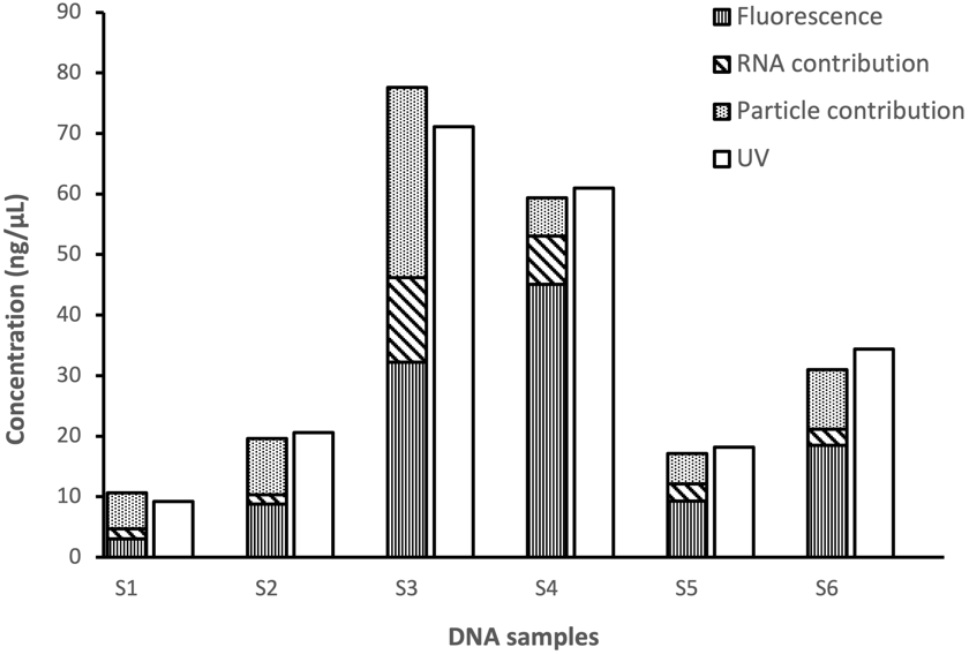
Contribution of magnetic particles and RNA to DNA concentration measurements by UV absorbance spectrophotometry.

## Discussion

In the present study, we showed that residual magnetic particles used for lineage-specific cell isolation were present in the corresponding genomic DNA samples, and that these particles interfered with DNA quantification by UV spectrophotometry. The amount of particles carried through the DNA extraction column was variable, and was probably influenced by the column physical properties, as well as the composition and viscosity of the cell lysate. On average, removing the particles from the DNA samples decreased the measured concentration by 37%. When combined with the presence of RNA, the DNA concentration as measured by UV spectrophotometry was roughly twice as high as the actual concentration. Different degrees of DNA concentration overestimation would likely be observed with magnetic particles and DNA extraction methods different from the ones used in the present study.

Because of its convenience, UV spectrophotometry will likely remain the DNA quantification method chosen by many laboratories. However, during the validation phase of a new lineage-specific DNA isolation workflow, DNA samples concentrations should be assessed by both UV spectrophotometry and fluorescence. This would provide an estimate of the effect of particles and RNA on UV measurements, which should be taken into account when adjusting DNA concentration for downstream assays.

## Supporting information

Dat supplements Table 1

## Data Availability

All data produced in the present study are available upon reasonable request to the authors.

## Notes

### Competing Interest Statement

The authors have declared no competing interest.

### Funding Statement

Izza Usman Bajwa was supported by grants from BioAlberta and BioTalent Canada.

### Author Declarations

The Institutional Review Board of WCG IRB gave ethical approval for the collection and use for research of the blood samples used for this work.

