## Supplementary material for "Effect of cell isolation magnetic particles on DNA quantification by UV absorbance spectrophotometry": Dat supplements Table 1

| DNA sample | [DNA] measured by UV spectrophotometry (ng/μL) | | A260/A280 | | A260/A230 | | [DNA] measured by fluorescence (ng/μL) | | [RNA] (ng/μL) |
|---|---|---|---|---|---|---|---|---|---|
| | After elution | After centrifugation | After elution | After centrifugation | After elution | After centrifugation | After elution | After centrifugation | After centrifugation |
| S1 | 9.2 | 3.3 | 2.17 | 3.77 | 1.00 | 1.34 | 3.2 | 3.0 | 1.4 |
| S2 | 20.6 | 11.3 | 1.99 | 2.64 | 0.93 | 1.09 | 9.6 | 8.8 | 1.2 |
| S3 | 71.1 | 39.7 | 1.81 | 1.91 | 1.14 | 1.52 | 31.6 | 32.2 | 11.2 |
| S4 | 61.0 | 54.7 | 1.86 | 2.00 | 1.59 | 2.14 | 43.6 | 45.1 | 6.4 |
| S5 | 18.2 | 13.2 | 1.64 | 1.81 | 1.31 | 2.01 | 11.3 | 9.3 | 2.3 |
| S6 | 34.4 | 24.6 | 1.69 | 1.76 | 1.09 | 1.81 | 16.1 | 18.5 | 2.1 |
